# SARS-CoV-2 neutralization after mRNA vaccination and variant breakthrough infection

**DOI:** 10.1101/2022.02.09.22270692

**Authors:** Christian Gaebler, Justin DaSilva, Eva Bednarski, Frauke Muecksch, Fabian Schmidt, Yiska Weisblum, Katrina G. Millard, Martina Turroja, Alice Cho, Zijun Wang, Marina Caskey, Michel C. Nussenzweig, Paul D. Bieniasz, Theodora Hatziioannou

## Abstract

Vaccination and infection by viral variants are shaping population immunity to SARS-CoV-2^1^ and breakthrough infections of vaccinated or previously infected individuals have become common as variants evade preexisting immunity. Omicron (B.1.1.529) is highly resistant to plasma neutralizing antibodies elicited by infection with prior variants and the 2-dose mRNA vaccination regimens. However, vaccination after infection or a third mRNA vaccine dose elicit high levels of neutralizing antibodies that can also neutralize omicron to a degree^2-4^. We compared neutralizing antibody titers in 54 individuals that had received 2 or 3 doses of mRNA vaccines and had experienced breakthrough infection with SARS-CoV-2 variants.

## Main text

We compared neutralizing antibody titers in 54 individuals that had received 2 or 3 doses of mRNA vaccines and had experienced breakthrough infection with SARS-CoV-2 variants. Breakthrough variants were deduced based on the prevalent variant circulating in New York City at the time of infection (Fig 1A, S1 and Supplementary appendix). Delta (B.1.617), was the predominant variant from July to early December 2021 but was rapidly displaced by omicron by January 2022 (Fig. 1A). As a consequence of evolving vaccination recommendations, 24 participants that experienced breakthrough infection with delta had received 2 vaccine doses. Conversely, most omicron breakthrough infections (24 out of 30) were in participants who received 3 vaccine doses.

**Figure-1.**
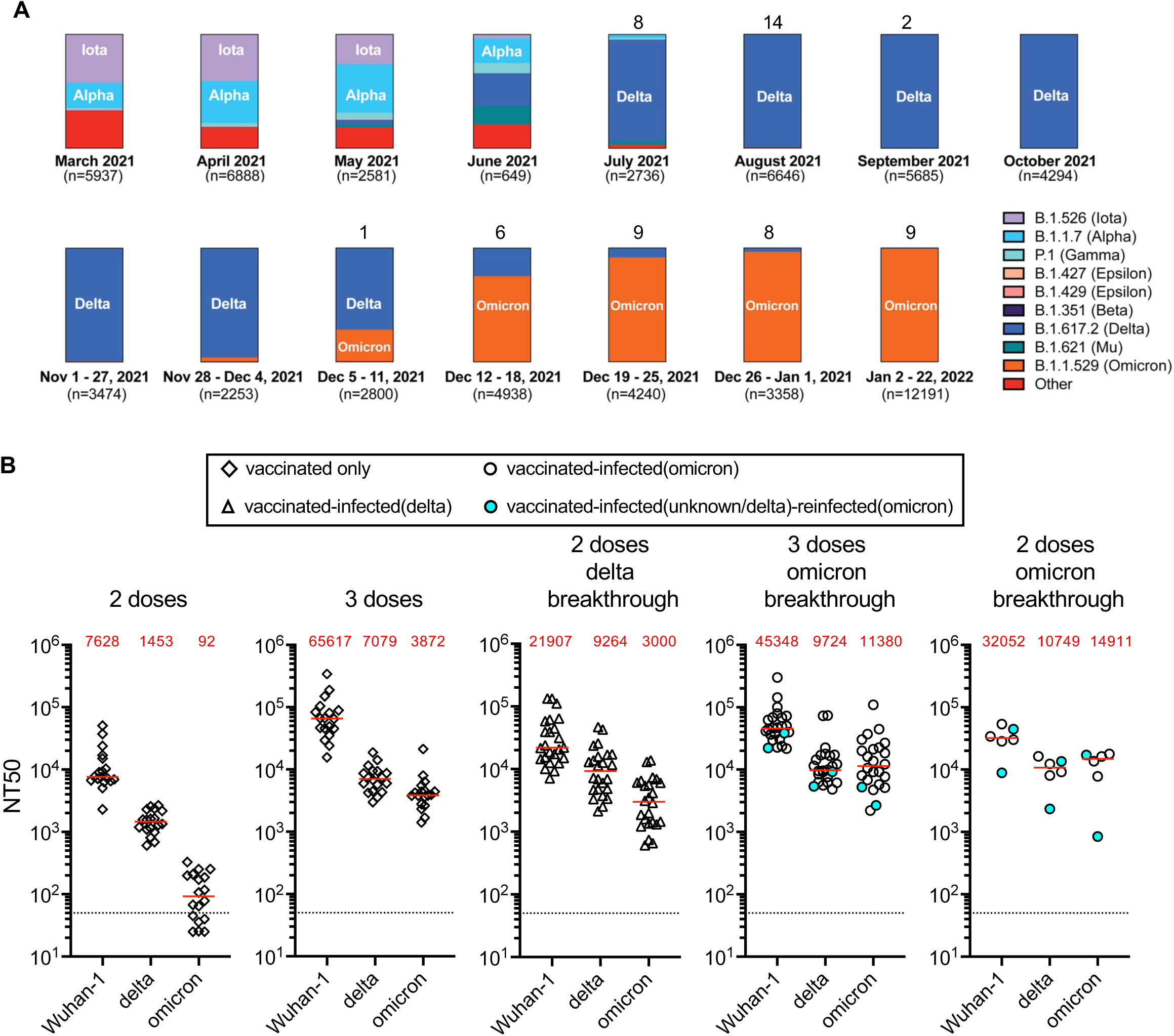
Variant prevalence and plasma neutralization titers. Panel A shows the frequency of variants over selected periods of weeks or months based on viral genome sequences obtained from samples collected in New York City and downloaded from NYC Department of Health and Mental Hygiene (https://github.com/nychealth/coronavirus-data/blob/master/variants/variant-epi-data.csv) on February 4, 2022. Numbers on top of bar charts indicate number of PCR positive samples identified over that period in our cohort. Panel B shows the NT50 values against Wuhan-1, delta (B.1.617) or omicron (B.1.1.529). Plasma samples were collected from uninfected, vaccinated individuals 29-53 days after the 2^nd^ or 3^rd^ dose of an mRNA vaccine (BNT162b2, Pfizer/mRNA-1283, Moderna) or from individuals that had been vaccinated with 2 or 3 doses of an mRNA vaccine and experienced one or two breakthrough infections with samples collected between 14-55 days after the latest positive test for infection. Horizontal lines and red values in each group indicated median NT50 values.

Compared to uninfected 2 dose vaccine recipients, participants with pre-omicron breakthrough infections had median plasma neutralization titers that were 2.8-, 4.9- and 26.4-times greater against Wuhan-hu-1, delta and omicron (all p≤0.0005) (Fig. 1B). Compared to 3 dose vaccine recipients, participants with omicron breakthrough had neutralizing titers that were 2.9-times greater (p<0.0001) against omicron and 1.4-times greater (p=0.0088) against delta and were not greater against Wuhan-hu-1 (Fig. 1B). For the few individuals with omicron breakthrough after only 2 vaccine doses, median neutralizing titers were 4.2-, 7.4- and 161.5- times greater against Wuhan-hu-1, delta and omicron (all p≤0.0074) than uninfected 2 dose recipients. Overall, in comparing vaccine doses and breakthrough infections, omicron breakthrough infection elicited the greatest increase in neutralizing antibody titers against omicron itself, but the magnitude of this effect was small in individuals that had received a third vaccine dose. Because omicron breakthrough infection broadened plasma neutralizing activity more effectively than delta variant infection (Fig. 1B), our data suggest that an omicron specific booster shot may have similar effects in those that received only 2 mRNA vaccine doses^5^. In individuals that have already received a third dose of the currently available mRNA vaccines, omicron specific boosters may modestly improve neutralizing responses against omicron while not significantly impacting neutralization of other variants.

## Data Availability

All data produced in the present work are contained in the manuscript

## Methods

### Study participants

Participants were recruited for blood donations at Rockefeller University Hospital in New York between August 13, 2021 and January 28, 2022. Eligible participants were healthy adults with a history of infection with SARS-CoV-2 after receiving two or three doses of Moderna (mRNA-1273) or Pfizer-BioNTech (BNT162b2) vaccines according to current dosing and interval guidelines. Clinical data collection and management were carried out using the software iRIS by iMedRIS (v.11.02). All participants provided written informed consent before participation in the study, and the study was conducted in accordance with Good Clinical Practice principles. The study was performed in compliance with all relevant ethical regulations, and the protocol (DRO-1006) for studies with human participants was approved by the institutional review board of The Rockefeller University. For detailed participant characteristics, see Supplementary Table 1.

Variants likely responsible for breakthrough infection in each participant were deduced based on the prevalent variants circulating in New York City at the time of infection diagnosis, except for one participant infected between December 5-11, where the contact suspected of having transmitted the virus to the study participant had a sequence-confirmed omicron infection.

Uninfected vaccine recipient participants have been previously desrcibed^1^.

### Blood sample processing and storage

Heparinized plasma and serum samples were aliquoted and stored at –20 °C or below. Before experiments, aliquots of plasma samples were heat inactivated (56 °C for 1 h) and then stored at 4 °C.

### Pseudotyped virus neutralization assays

SARS-CoV-2 pseudotyped particles were generated as previously described ^2^. Briefly, 293Tcells were transfected with pNL4-3ΔEnv-nanoluc and pSARS-CoV-2-SΔ19. At 48 hours later particles were harvested, filtered and stored at -80°C. The amino acid deletions and/or substitutions corresponding to SARS-CoV-2 variants were incorporated into a spike expression plasmid using synthetic gene fragments (IDT) or overlap extension PCR mediated mutagenesis and Gibson assembly. Specifically, the variant-specific deletions and substitutions introduced into the B1 sequence were:

B.1.617 (delta): T19R/Δ156-8/L452R/T478K/D614G/P681R/D950N

B.1.1.529 (omicron): A76V/Δ69-70/T95I/G214D/Δ143-145/Δ211/L212I/ins214EPE/G339D/S371L/S373P/S375F/K417N/N440K/G446S/S477N/T478K/E484A/Q493K/G498R/N501Y/Y505H/T547K/D614G/H655Y/N679K/P681H/N746K/D796Y/ N856K/Q954H/N969K/L981F

All spike proteins used in the pseudotype neutralization assays had a 19 amino acid C-terminal deletion and included the R683G substitution, which disrupts the furin cleavage site and increases particle infectivity without grossly affecting antibody sensitivity. Fivefold serially diluted plasmas from vaccinated individuals were incubated with SARS-CoV-2 pseudotyped virus for 1 h at 37 °C. The mixture was subsequently added to an HT1080-based cell line engineered to express human ACE2 (HT1080.ACE2 cl14). The starting serum dilution on cells was 1:50. Nanoluc Luciferase activity in lysates was measured 48 hours post-inoculation using the Nano-Glo Luciferase Assay System (Promega) with the Glomax Navigator (Promega). Relative luminescence units were normalized to those derived from cells infected with SARS-CoV-2 pseudotyped virus in the absence of serum. The half-maximal neutralization titers for sera (NT_50_) were determined using four-parameter nonlinear regression (least squares regression method without weighting; constraints: top=1, bottom=0) (GraphPad Prism) and median values calculated for each sample from 2-3 independent experiments. Group comparisons were performed using Mann-Whitney test (GraphPad Prism).

The time between diagnosis of infection and sample collection did not significantly affect NT50 values (Fig. S1).

## Acknowledgements

We are grateful to all participants that volunteered for this study. We thank M. Bergh, M. Okawa Frank and R. B. Darnell for the occupational SARS-CoV-2 infection surveillance program and participant referrals. Supported by grants from the National Institutes of Health (R01AI501111, to Dr. Bieniasz; R01AI78788, to Dr. Hatziioannou; P01-AI138398-S1 and 2U19AI111825, to Dr. Nussenzweig and P01AI65075 to Drs Bieniasz, Hatziioannou and Nussenzweig). Dr. Gaebler’s work is supported by the Robert S. Wennett Post-Doctoral Fellowship, the National Center for Advancing Translational Sciences (National Institutes of Health Clinical and Translational Science Award program, grant UL1 TR001866), and the Shapiro–Silverberg Fund for the Advancement of Translational Research. Drs. Bieniasz and Nussenzweig are Howard Hughes Medical Institute Investigators (HHMI). This article is subject to HHMI’s Open Access to Publications policy. HHMI lab heads have previously granted a nonexclusive CC BY 4.0 license to the public and a sublicensable license to HHMI in their research articles. Pursuant to those licenses, the author-accepted manuscript of this article can be made freely available under a CC BY 4.0 license immediately upon publication.

## Author contributions

C.G., M.C.N., P.D.B. and T.H. conceived, designed and analyzed the experiments. J.D.S, E.B, F.M., F.S., Y.W., and A.C. performed pseudotype neutralization experiments. C.G., Z.W., K.G.M., M.T. and M.C. executed clinical protocols and recruited participants and collected and processed samples. C.G., P.D.B., M.C.N. and T.H. wrote the manuscript.

## Supplementary figure legends

**Figure S1.**
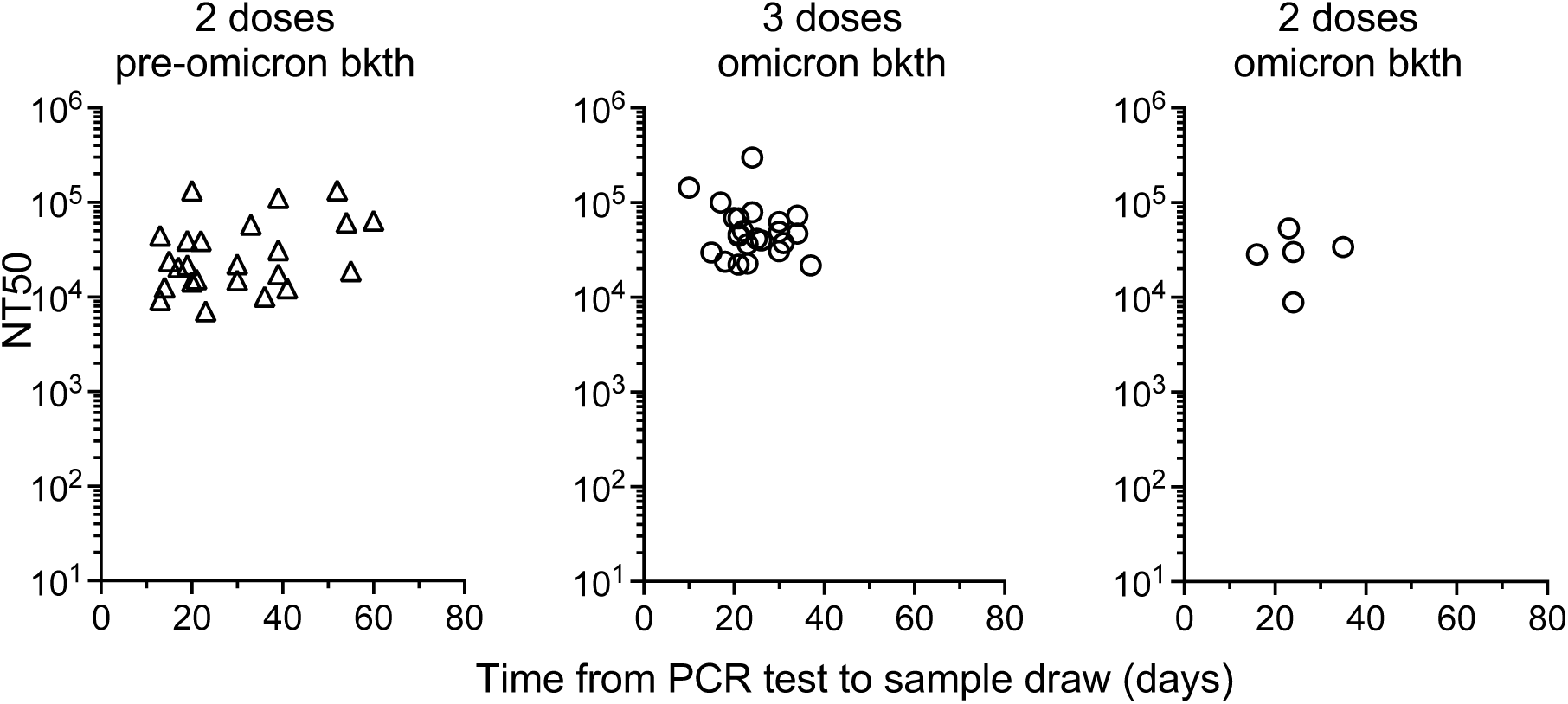
Correlation of time after sample collection and neutralizing antibody titers. The NT50 values from samples collected from vaccinated individuals that experienced a breakthrough infection is shown relative to the time (days) between infection diagnosis and sample collection.

**Table S1.**
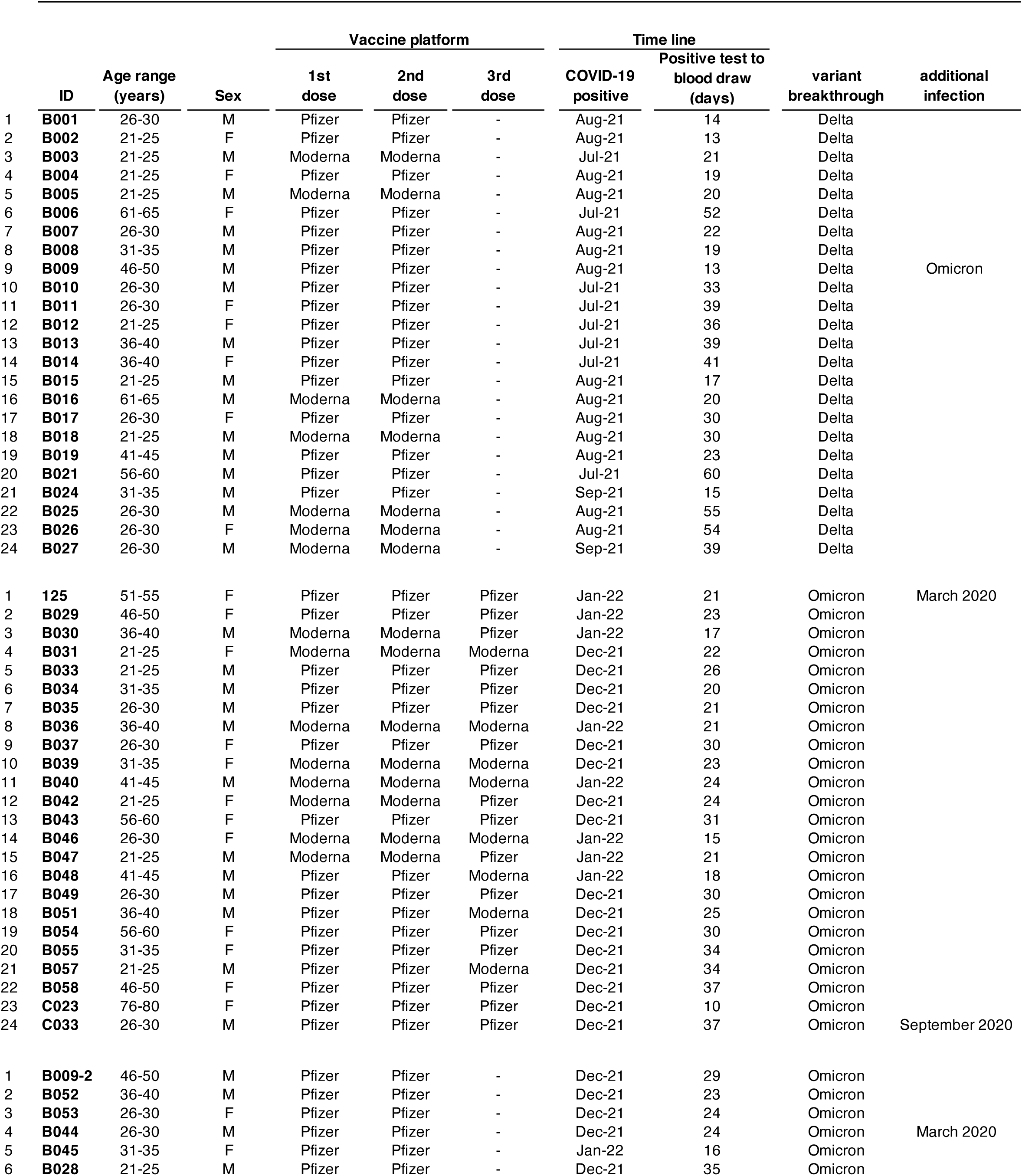
Individual participant characteristics.

